# The final size of an epidemic in a heterogeneous society

**DOI:** 10.1101/2022.10.10.22280914

**Authors:** Peter de Jong

## Abstract

This note describes the outcome of an epidemic in a heterogeneous population with a very simple structure. The population is split into two sections in which the epidemic runs a different course. The reproduction numbers of the two epidemics are unobserved, only the overall reproduction number is known. For such a population the outcome of the epidemic can be as well far worse as far better than expected on the basis of the overall reproduction number.

By considering a very simple model of this population some calculations are feasible under general assumptions on the epidemic itself. These calculations show in which direction models based on the overall reproduction number can misrepresent the real-world situation.

## Introduction

This note is concerned with an epidemic in a heterogeneous population with a very simple structure. The population of size *n* is divided into two parts with sizes *αn* and (1 − *α*)*n*. The epidemic has distinct reproduction numbers in each part of the population and there are no cross infections. The reproduction numbers, *R* and *R*′ are not observed; only the weighted average *R*_0_ = *α R* + (1 − *α*)*R*′ is observed. So the model depends on two parameters *R*_0_ and *α*.

As a proxy for the impact of the epidemic we take the final fraction, that is the fraction of the population infected during the course of the epidemic. We compare the impact of the epidemic in a structured population to that in a homogeneous population with reproduction number *R*_0_. We take the ratio of both final fractions and thus the ratio of the total numbers of infected individuals in both cases.

Since both *R* and *R*′ are not observed, we consider the curve of the ratio as a function of *R* in the range [0, *R*_0_/*α*], representing all possible outcomes for given values of *R*_0_ and *α*. The curve has different characteristics for different values of *R*_0_. Broadly speaking, for small values of *R*_0_ (we take always *R*_0_ >1) the outcome of an epidemic in our structured population can be far more severe than in a homogeneous population with the same reproduction number. For higher values of *R*_0_ this phenomenon reverses and any partition of a population into disjoint parts with different *R*-values dampens the effect of the epidemic.

Obviously, in order to make these rather general observations some assumptions are necessary. In fact, all results in this note rest on one assumption regarding the form of the graph describing the final fraction as function of the reproduction number. Any epidemic model that produces a final fraction curve of this form will fit into our results. There is a broad class of models that produce final fraction curves with the desired characteristics. Any epidemic model that satisfies the final size equation falls into this class. Very different models satisfy this equation, ranging from the simple case of the Reed–Frost model (see (Britton 2018)) to models involving Erdös–Rényi networks and models allowing super-spreaders, as is shown in (Ma and Earn 2006) and (Miller 2012).

There is an unusual aspect in our model: a curious lack of information. The reproduction numbers in the separate parts are not known and it is assumed that the overall reproduction number *R*_0_ is the weighted average of the unknown *R*-numbers. With more specific information a different model would have applied. One might say that our model describes the possible outcomes in the presence of a special ‘known unknown’, the information whether an individual belongs to one group or the other. Without this information it is impossible to determine the *R*-numbers per subpopulation and the overall *R*_0_ will be measured as the weighted average of the *R*-numbers in the subpopulations, assuming both groups are sampled equally.

In many societies deep divides exist between groups along lines of ethnicity or religion. Often these divisions coincide with differences in working conditions, housing, family size, and access to health care and adequate information. These groups are likely to be affected differently by epidemics (and many other health issues), resulting in distinct reproduction numbers. Some national healthcare systems have in varying degree access to data relating to these kinds of group membership. For example the UK has a modern wide ranging healthcare information system encompassing data on race. For some of the results based on it see (Morales and Ali 2021) and (Mathur et al. 2021), also indicating the limits on the information available. For reflection on the historical limits of these data see (Morgan 2020).

Indigenous people form a special case, where some national healthcare systems spend extra effort to gather specific information and provide focused care. See (Connolly, Griffiths, et al. 2021) for a very recent overview and for the situation in some specific countries see (Connolly, Jacobs, and Notzon 2021) and (Griffiths et al. 2021).

Many countries do not record ethnicity or religion out of historical concerns or privacy considerations. Estimates of the size of these groups, that is estimates of *α*, are available via surveys. Variables that describe group membership on the level of individuals are not available, at least not in the data from which the reproduction number is estimated. These data appear to come from a homogeneous population. This is the setting of the model that will be analyzed in the next section.

## Analysis of the ratio curve

Consider a population of size *n* that is partitioned into two parts of size *αn* and (1 − *α*)*n* with separate epidemics in the two parts. In order to avoid side effects caused by too small numbers *αn* or (1 − *α*)*n* the parameter *α* is taken sufficiently away from 0 and 1, 0 *≪ α ≪* 1.

There are no cross infections, the two epidemics are completely isolated with reproduction numbers *R* and *R*′. The model discussed in this note is characterized by the linear relation *αR* + (1 − *α*)*R*′ = *R*_0_ which will be the reproduction number measured when sampling is conducted without information on the partitioning.

Let *Z* be the final fraction of the population, that is the fraction infected during the course of the epidemic. Here *Z* = *Z*(*R*) is taken as a function of *R* and is the indicator for the severity of the epidemic. We will consider final fraction curves of a special form.

### Definition 1.

*The function Z* : [0, ∞) → [0, 1], *Z* ≢ 0, *is called (strictly) QC if Z*(*R*) = 0 *for* 0 ≤ *R* ≤ 1 *and Z is (strictly) concave for R* ≥ 1.

### Remark 1.

The trivial function *Z* ≡ 0 is excluded to avoid trivial problems.

### Remark 2.

If *Z* is (strictly) QC, then *Z* is (strictly) increasing on (1, ∞).

The class of epidemics with final fraction curves that are QC is not empty. It contains in any case the models satisfying the final size equation,

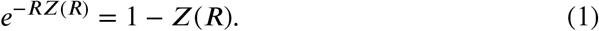

### Remark 3.

Clearly, the trivial solution *Z* ≡ 0 satisfies the final size equation.

### Lemma 1.

*Functions* ***Z***(***R***) ≢ 0 *satisfying the final size equation are strictly QC*.

*Proof*. For 0 ≤ *R*≤ 1 the only solution is *Z*(*R*) = 0, since, for *Z*(*R*) >0 we have *e*^−*RZ*(*R*)^ >*e*^−*Z*(*R*)^ > 1 − *Z*(*R*). The solution *Z*(*R*) ≢ 0 is concave for *R* ≥ 1 since the inverse: *R* = − log(1 − *Z*)/*Z*, with 0 *< Z <* 1 is striclty convex as can be seen from the series expansion of the inverse. □

It is assumed that in the two parts of the population the epidemic has the same final fraction curve, possibly with different reproduction numbers. This note is concerned with the ratio of the sum of the final sizes in both parts and the final size of the epidemic in the undivided population with reproduction number *R*_0_, the weighted average of the reproduction numbers in the parts. The ratio is taken as function of *R*, the reproduction number in one part with the range [0, *R*_0_/*α*], displaying for fixed *R*_0_ and *α* all possible outcomes of the epidemic.

### Definition 2.

*Let Z be QC as in Definition 1. Then define for fixed α*, 0 *≪ α ≪* 1, *and R*_0_ >1 *the ratio curve g* : [0, *R*_0_/*α*] → [0, 1) *by*

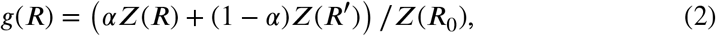

*with R and R*′ *satisfying the linear relation αR* + (1 − *α*)*R*′ = *R*_0_, *or R*′ = (*R*_0_ − *αR*)/(1 − *α*).

Below the ratio curve *g* is described in terms of extrema with *g* monotonous in between. The locations of the extrema only depend on *R*_0_ and *α*, not on the specific form of *Z*, as long as *Z* is QC.

### Theorem 1.

*The function g, defined in Definition 2, has maxima at R* = 0, *R* = *R*_0_ *and R* = *R*_0_/*α. Furthermore g has minima at R* = 1 *and R*′ = 1, *that is at R* = *R*_0_/*α* − (1 − *α*)/*α*.

*If the function Z*(*R*) *is strictly QC, then g*(*x*) *is strictly monotonous between extrema*.

*Proof*. The proof consists in showing that *g*(*R*) is (strictly) monotonous between extrema, with the correct direction. In the proof it is assumed that *Z*(*R*) is strictly QC.

For *R* ≤ 1 we have *Z*(*R*) = 0 and *g*(*R*) reduces to *g*(*R*) = (1 − *α*)*Z*(*R*′) which is strictly decreasing since *Z* is an increasing and *R*′ is a decreasing function of *R*. In the same way, for *R ≥ R*_0_/*α −* (1 − *α*)/*α*, that is for *R*′ ≤ 1 the resulting *g*(*R*) = *α Z*(*R*) is increasing.

If 1 *< R < S < R*_0_ then *R*_0_ *< S*′*< R*′ and since *Z* is strictly concave, the line through (*S, Z*(*S*)) and (*S*′, *Z*(*S*′)) is strictly above the line through (*R, Z*(*R*)) and (*R*′, *Z*(*R*′)) (see for example Rudin 1987), which implies *g*(*R*) *< g*(*S*) and hence *g* is increasing on the interval (1, *R*_0_). In the same way it is shown that *g* is decreasing on the interval (*R*_0_, *R*_0_/*α −*(1 − *α*)/*α* that is *R*_0_ >*R*′ >1. This proves the theorem. □

The extrema given in the above theorem depend on the value *R*_0_, except the local maximum at *R* = *R*_0_ which obviously equals 1 for all values of *R*_0_. In general one can say that extrema increase with diminishing *R*_0_ and diminish with increasing *R*_0_. The next theorem shows how the extrema of the ratio curve behave for values of *R*_0_ close to 1 and for sufficiently large *R*_0_.

### Theorem 2.

*Let Z be QC and suppose the right-hand derivative of Z at R* = 1 *exists and is finite, then we have the following limits for the extrema*.

| Location of the extremum $R$ | $g(R)$ | $\lim_{R_0 \downarrow 1} g(R)$ | $\lim_{R_0 \rightarrow \infty} g(R)$ |
| --- | --- | --- | --- |
| 0 | $\frac{Z(R_0/(1-\alpha))}{Z(R_0)/(1-\alpha)}$ | ∞ | $1-\alpha$ |
| 1 | $\frac{Z((R_0-\alpha)/(1-\alpha))}{Z(R_0)/(1-\alpha)}$ | 1 | $1-\alpha$ |
| $R_0$ | 1 | 1 | 1 |
| $(R_0 - (1-\alpha))/\alpha$ | $\frac{Z((R_0 - (1-\alpha))/\alpha)}{Z(R_0)/\alpha}$ | 1 | $\alpha$ |
| $R_0/\alpha$ | $Z(R_0/\alpha)/(Z(R_0)/\alpha)$ | ∞ | $\alpha$ |

### Remark 4.

The extra condition on the righthand derivative of *Z*(*R*) is only required to prove the limits for the minima for *R*_0_ ↓ 1.

*Proof*. The extremum at *R* = *R*_0_ needs no further treatment since it equals 1 for all values of *R*_0_. The remaining maxima have the form *Z*(*βR*_0_)/*βZ*(*R*_0_) with *β* = 1/(1 − *α*) and *β* = 1/*α*, the minima have the form *Z*(*β*(*R*_0_ − 1) + 1)/*β Z*(*R*_0_) again with *β* = 1/(1 − *α*) and *β* = 1/*α*.

The limits for *R*_0_ → ∞ follow from the fact that *Z*(*R*) has a finite, non-zero limit for *R* increasing: Then we have

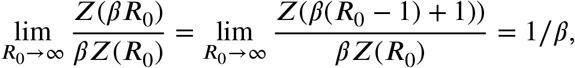

which results in the limits in the rightmost column.

Since *Z*(*β*) >0 and since 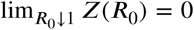 the limits for the maxima at *R* = 0 and at *R* = *R*_0_/*α* follow.

The limits for the minima can be written as:

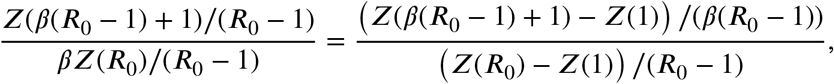

and, if the right-hand derivative of *Z* exists and is finite, the limits for the minima follow. □

The above theorem applies to the extrema of *g* for limit values of *R*_0_. Some general remarks can be made. As long as the epidemic is growing in both parts of the population, that is *R* >1 and *R*′ >1, for all values of *R*_0_ the partitioned population does not suffer a worse outcome than the homogeneous population, in our model: *g* ≤ 1. In many instances the outcome may be better than expected, *g* < 1.

This is, however, not the case with two really different epidemics in the population: *R <* 1 or *R*′ *<* 1. Low values of *R*_0_ can lead to outcomes that are worse than projected on the basis of, at first sight, sound observations. The exact values of *R* and *R*_0_ for which this worrying situation occurs, depend on the specific form of *Z*(*R*).

## Discussion

A first step towards more realistic epidemic models is the introduction of population heterogeneity. The structure of the population in this note is very simple, a bipartite population without cross-infections between the parts. This simplicity allows us to show in some detail the effects of heterogeneity parametrized by a single parameter, *R*_0_.

A more accurate model for population heterogeneity is age structure with transmission rates within and between age groups. This yields a ‘next generation matrix’ which can be used for modeling, see for example (Britton, Ball, and Trapman 2020). Transmission rates depend on quite sophisticated data about social interactions between and within age groups. In this more sophisticated model the issue of divisions in a society resurfaces in a more complex and intractable form. The simple model of one partitioned population is replaced by multiple partitioned fractions and their interactions. The research that produced sociometric data underlying (Britton, Ball, and Trapman 2020), extensive surveys in two middle-sized Dutch cities in 1986, hints in this direction. The survey was multilingual and respondents were, if necessary, approached in other languages than Dutch (Wallinga, Teunis, and Kretzschmar 2006).

From a more general point of view this note shows, once more, that averaging can lead to misleading results. However, since in this case averaging follows directly from a lack of information on the level of individual observations, it will be hard to avoid. Therefore models that rely critically on measurements of *R*_0_ should be treated with some care.

## Data Availability

All data produced in the present work are contained in the manuscript

## Appendix

The above results are illustrated by a few plots of the ratio curve for different values of *R*_0_. The shape of the ratio curve depends on the two parameters *α* and *R*_0_ and on the shape of the final fraction curve. The plots below are for *α* = 1/3.

As final fraction curve is taken the solution of the final size equation (1). The solution cannot be expressed in terms of elementary functions, but it can be expressed in terms of the Lambert W function. Thus, as final fraction curve is taken the following expression:

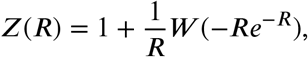

with *W* (*x*) the unique solution of the transcendental equation 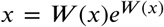. Here *W* is the principal branch of the Lambert-W function, for 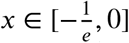, see appendix A in Ma and Earn 2006. Here we use the function `lambertW0` from the `lamW`-package in R.

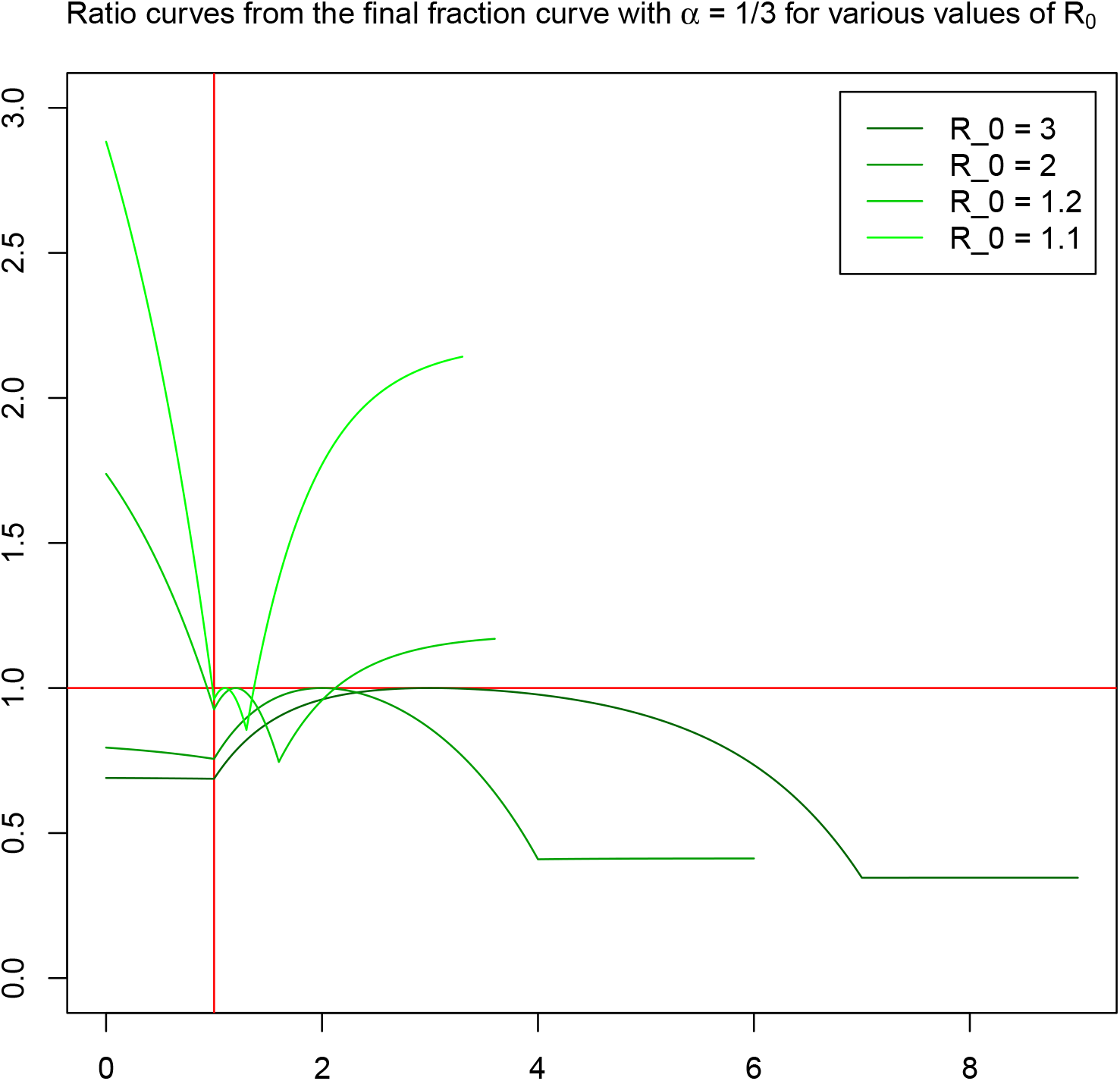

Note that the plots for different *R*_0_ are defined on different ranges [0, *R*_0_/*α*]. Trivially the ratio curves equal 1 in *R*_0_. The above plots illustrate the general characteristics of the ratio curve: for small values of *R*_0_ the final fraction can be above 1, indicating an worse outcome in a structured population than in a homogeneous population with the same *R*_0_. And for higher values of *R*_0_ the revers holds: any partition of the population into disjoint parts with distinct reproduction numbers will reduce the impact of the epidemic with a ratio curve below 1.

